# Disruption of outdoor activities caused by wildfires increases disease circulation

**DOI:** 10.1101/2024.08.08.24311678

**Authors:** Beatriz Arregui-Garcıa, Claudio Ascione, Arianna Pera, Boxuan Wang, Davide Stocco, Colin J. Carlson, Shweta Bansal, Eugenio Valdano, Giulia Pullano

**Author notes:** These authors contributed equally to this work (Beatriz Arregui-Garcıa, Claudio Ascione, Arianna Pera).

## Abstract

Although climate change poses a well-established risk to human health, present-day health impacts, particularly those resulting from climate-induced behavioral changes, are under-quantified. Analyzing the U.S. West Coast wildfires of September 2020, we found that poor air quality drives people indoors, increasing the circulation of airborne pathogens like COVID-19. Indoor masking rates as low as 10% can mitigate this risk, offering a clear path to enhance public health responses during wildfires.

## Main

Climate change is significantly increasing the frequency and intensity of extreme weather events, posing severe threats to both the environment and human health. Heatwaves, storms, floods, and wildfires not only devastate ecosystems and economies but also pose significant risks to human health [1]. These extreme events lead to significant loss of life, disability, and well-being through a mix of injuries, infectious diseases, and non-communicable diseases [2].

Extreme weather events also influence human behavior [3], which in turn might affect human health indirectly [4]. This has the potential to create compound risks for human health during ongoing public health emergencies [1]: for example, during the first three years of the Covid-19 pandemic, unprecedented heatwaves drove many people indoors, leading to a substantial number of excess Covid-19 cases and deaths [5-6]. However, the cascade effects of localized or permanent behavioral changes due to extreme weather events on the dynamics of infectious diseases remain underexplored. Addressing this gap is essential for enhancing our ability to predict and effectively respond to future epidemics in a world that is already 1.3°C warmer.

Wildfires are among the most disruptive events for human behavior, severely impacting air quality across vast regions [7] and posing significant concerns for public health. While much of the public health focus has been on the physiological effects of wildfires [8]—such as direct mortality due to short-term exposure to fine particulate matter (PM_2.5_) air pollution [9], and synergistic effects of PM_2.5_ exposure on respiratory infectious disease susceptibility and severity [10]—there has been less attention to how wildfire-induced changes in behavior might influence the spread of airborne diseases. However, perceptions of deteriorating air quality and government advisories [11] often lead individuals to reduce outdoor activities. This shift results in increased indoor activity, where conditions such as close social proximity, limited ventilation, and the accumulation of respiratory droplets can facilitate the transmission of airborne diseases like influenza and SARS-CoV-2 [12-13]. From a public health perspective, with the increasing frequency and severity of wildfires, it has become crucial to understand how disruptions in outdoor activities during wildfires might indirectly impact disease dynamics in indoor settings.

To address this question, we focused on the 2020 wildfire season, one of the most intense in recent decades. In August 2020, thunderstorms ignited multiple wildfires across California, Oregon, and Washington [14]. To characterize disruptions in human behavior and their impact on pathogen circulation, we used a weekly time series of the relative tendency of human interactions to occur indoors versus outdoors in U.S. counties. This metric, called the indoor activity seasonality index, focuses on U.S. counties with unhealthy Air Quality Index (AQI) levels during the 2020 wildfire season in the Western United States [15]. By systematically quantifying the impact of poor air quality on seasonal outdoor activities and the indirect effect on airborne disease dynamics, we aim to better understand these relationships and enhance public health strategies in response to wildfires.

During and immediately following the wildfire event on September 10, 2020, multiple U.S. counties in Oregon and Washington reported significant AQI spikes. We focused on ten counties that reported an AQI exceeding 150 for at least three consecutive days post-wildfire, namely affected counties. We refer to counties with AQI < 100 as unaffected counties. As shown in Figure 1, our analysis revealed a significant increase in indoor activity in all affected counties, expect for Marion County, Oregon. Using a regression discontinuity approach [16], we find both Washington and Oregon saw in average an 11.6% and 13.0% increase in indoor activity, respectively, for about two weeks following the wildfires (Table S1).

**Figure 1:**
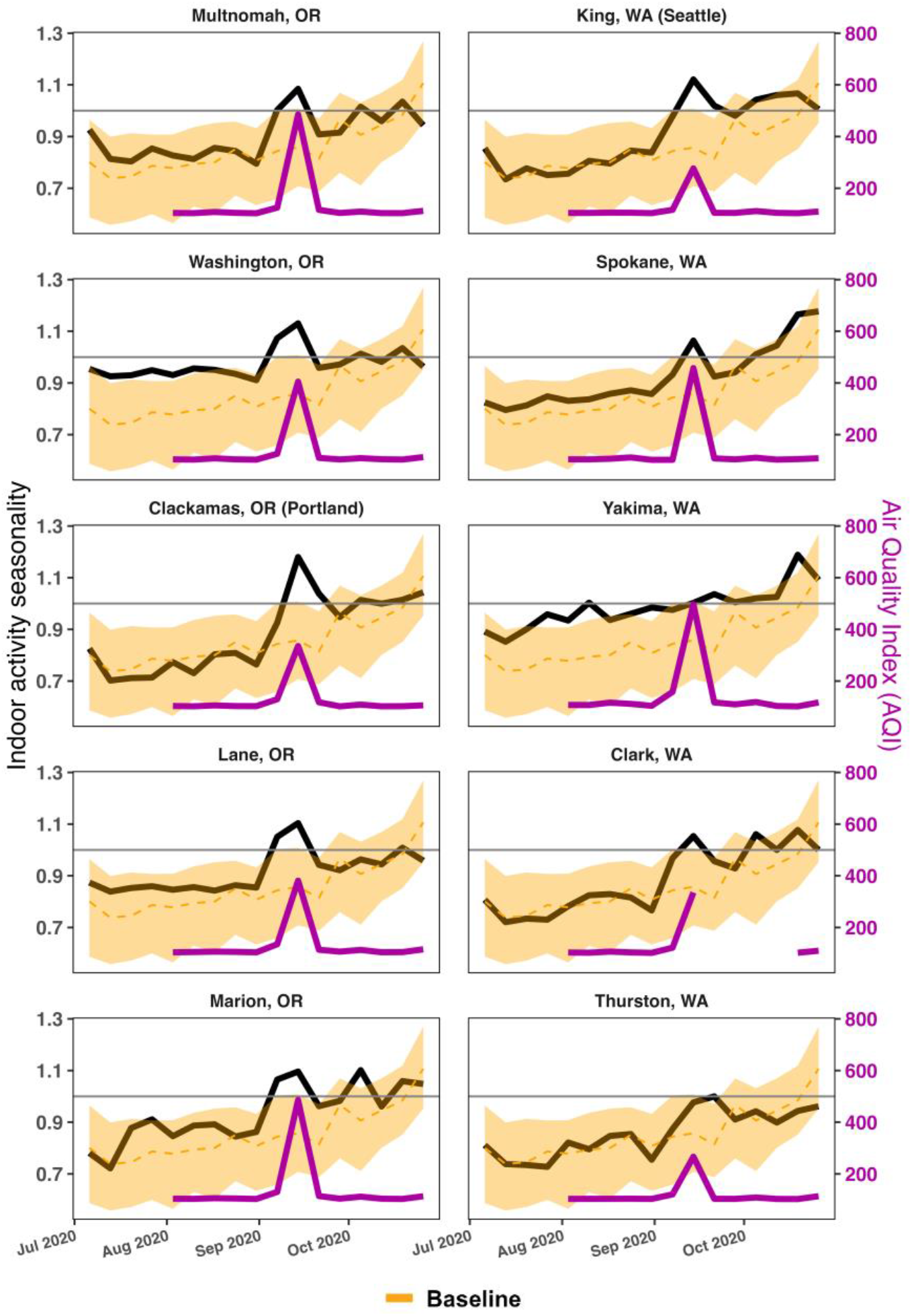
Atypical shift in human behaviors due to wildfire smoke. The figure shows the indoor activity seasonality index between July 1, 2020, and November, 01, 2020 in the 10 selected affected counties: Multnomah County, Washington County (Portland city), Clackamas County, Lane County, Marion County in Oregon state, and King County (Seattle city), Spokane County, Yakima County, Clark County, Thurston County in Washington state. In each subplot, we also show in yellow the median and 95% CI of the indoor activity seasonality index for the unaffected counties that we used as a baseline. The violet curves represent the AQI of the affected counties during the studied period.

We then studied the impact on respiratory pathogen circulation before, during, and after the AQI alert in the selected counties. Embedding the indoor index seasonality into an infectious disease transmission model, we find the impact of increased indoor activities on disease spread diminished with longer generation times of pathogens, i.e., the average time between the infection of a primary case and one of their secondary cases (Figure 2A). We also test different reproduction ratios (*R*_0_) consistent with seasonal respiratory viruses and currently circulating strains of SARS-CoV-2 (Figure 2B). Findings indicated that peak incidence in wildfire affected counties surpassed that of unaffected counties, with the highest relative peak incidences in Washington County, OR (+1.98), and Yakima County, WA (+1.65). The impact of increased indoor activities on disease spread diminished with higher *R*_0_. Similar results are found by looking at the relative attack rate (see SI).

**Figure 2:**
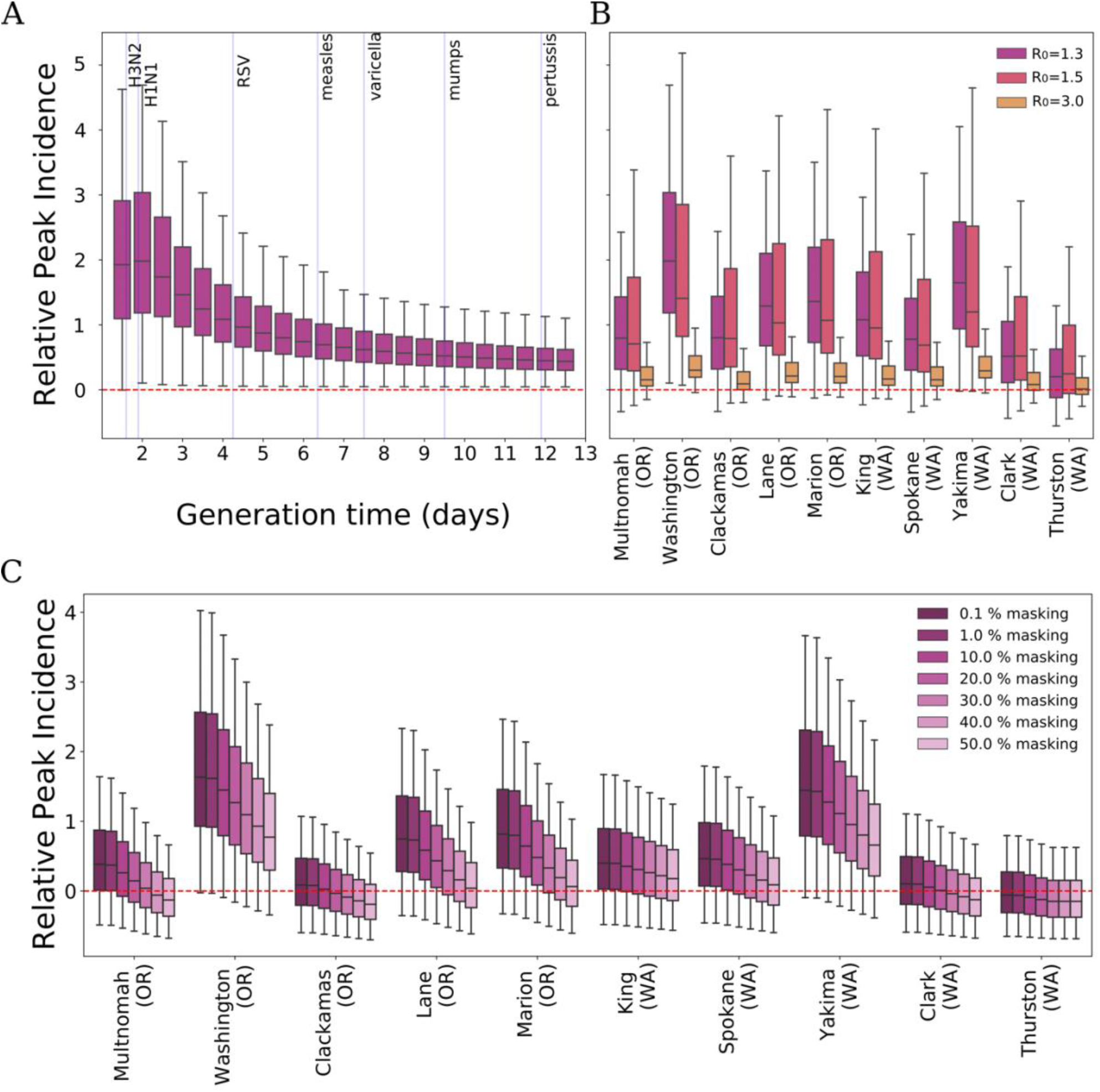
The cascade impact of the shift in human behavior on airborne diseases. A) Exploration of the relative variation in peak incidence (Washington County, OR) for different values of the generation time [19] with *R*_0_ = 1.3. Blue vertical lines indicate the average generation time of different airborne respiratory diseases. B) Relative variation in peak incidence for the selected 10 affected counties compared to the unaffected ones for three different reproduction ratios *R*_0_ = 1.3 (flu-like w/ interventions), *R*_0_ = 1.5 (flu-like). and, *R*_0_ = 3.0 (COVID-19-like). C) Relative variation in peak incidence when masking interventions are put in place, with *R*_0_ = 1.3.

To mitigate the increased exposure risk during wildfires, we tested the role of masking in indoor settings for several fractions of the population wearing masks. An increase in mask usage among the affected population significantly mitigated the rise in peak incidence caused by increased indoor activities (Figure 2C). These findings suggest that indoor masking should be strongly recommended during air quality alerts to counter the higher risk of virus exposure due to prolonged and crowded indoor activities.

This work relies solely on the ratio of indoor-to-outdoor activities, neglecting indoor activities at home, which could also impact epidemic circulation. Nevertheless, our primary objective is to elucidate the relationship between shifts in indoor behavior and respiratory disease spread in crowded public settings, to inform targeted interventions. Additionally, our model does not incorporate public health data, underscoring the urgency for further research utilizing such data to explore climate change implications for specific diseases more comprehensively.

Our approach highlights the need for further research into the impact of short-term declines in air quality caused by wildfire smoke on the transmission of respiratory diseases. The significant change in human behavior during wildfires observed across various U.S. states suggests that a targeted non-pharmaceutical intervention – indoor masking – could offset the increased risk of exposure. This study underscores the importance of tracking human behavior during and after extreme events to quantify how disruptions may affect pathogen exposure, increasing the probability of new outbreaks. Moreover, it emphasizes how public health responses to climate change must consider behavioral shifts induced by extreme weather events.

## Methods

Our study focuses on the 2020 wildfire season in the Western United States, specifically targeting the severe wildfire event from September 10 to November 1, 2020. During this period, we tracked air quality data from the U.S. Environmental Protection Agency (EPA) to identify counties where the AQI exceeded 150 for at least three days, indicating unhealthy air conditions [17]. We then selected the top five most populous counties, resulting in a focus on 10 counties impacted by wildfire smoke. This selection was refined using indoor activity data to ensure consistency in seasonal behavior patterns across selected US counties [13].

Our analysis proceeded in two steps. First, we characterize weekly human behavior patterns during the wildfire event in both affected and unaffected areas to identify significant behavioral shifts. Second, we integrated these behavioral changes into a mechanistic model to assess their impact on airborne diseases.

To compare affected and unaffected counties, we used the daily AQI, designating counties with AQI levels below 100 (Good or Moderate) during the wildfire event as unaffected. We selected 50 unaffected counties with populations in the top 25% of the population distribution to serve as a baseline for our analyses.

Human behavior was quantified using the indoor seasonality index by county [13], which tracks the tendency of individuals to engage in indoor versus outdoor activities. This index, derived from data collected from 2018 to 2021, measures activity at over 4.6 million points of interest, excluding home locations.

To identify discontinuities in indoor activity patterns, we employed a regression discontinuity (RD) approach [16]. The RD method assumes that indoor activity seasonality remains consistent over the study period, except for exposure to wildfire events. Using a local linear regression model, we estimate the event’s effect by fitting separate regression lines to observations before and after the event. For additional details on the RD method, please refer to the Supplementary Material (SI).

Next, we assessed the impact of behavioral shifts on epidemic circulation using a Susceptible-Infected-Recovered (SIR) model. This deterministic compartmental model simulated the spread of respiratory pathogens from seven days before the AQI alert to 14 days after. We calculated the relative variation in disease incidence by comparing the modeled peak incidence in affected counties with that in unaffected counties.

The model explores various reproduction numbers (*R*_0_) and generation times to account for different airborne infectious diseases. In addition, several masking scenarios were incorporated into the model, where we assume that individuals who wore masks reduced their susceptibility by 60% [18]. We studied the effect on disease incidence at the peak and on the overall attack rate.

Our findings revealed that wildfire-induced deterioration in air quality significantly increased indoor activities, which-in our simulations-elevated the spread of respiratory diseases. The study highlighted the importance of targeted interventions, such as indoor masking, to mitigate health risks during wildfire events. This research underscores the need for comprehensive public health strategies that consider behavioral shifts induced by extreme weather events and their implications for disease transmission.

## Data Availability

The data generated in this study are publicly available in the GitHub repository (https://github.com/GiuliaPullano/wildfires_project). The daily Air Quality Index (AQI) in all U.S. counties is collected by the Environmental Protection Agency (EPA) and is available on their website.

## Code availability

The code used in the study to analyze the shifts in behavior and the epidemic model developed to quantify the impact of infectious disease dynamics are archived and publicly available in the GitHub repository: https://github.com/GiuliaPullano/wildfires_project.

## Funding

Research reported in this publication was supported by the National Institutes of Health under award number R01GM123007.

## Contributions

SB, EV and GP conceived of and designed the study. GP, BA, CA, AP, DS, BW analyzed the data and did the analysis. BA implemented the model and ran the simulations. CC, SB, EV and GP interpreted the results. GP drafted the manuscript. All authors contributed to the writing of the final version of the manuscript.

## Acknowledgments

We acknowledge the Complexity72h workshop, held at IFISC in Palma, Spain, 26-30 June 2023, where this study was conceived. We also thank the Fritz-Family fellowship program to support the work.

## Notes

### Competing Interest Statement

The authors have declared no competing interest.

### Author Declarations

The indoor seasonality index we used is publicly available at https://github.com/bansallab/indoor_outdoor.

### Summary of Updates

Table S1 and one reference.

