## Supplementary Information for "Disruption of outdoor activities caused by wildfires increases disease circulation"

### County selection

We selected counties in Oregon (OR) and Washington (WA) reporting an Air Quality Index (AQI) greater than 150—indicating unhealthy air—for at least three days between July 1, 2020, and November 1, 2020. To avoid misleading effects of different seasonal behaviors between counties, we then filtered only those counties in the "Northern" indoor activity cluster as defined in [1]. For each state, we considered the top five counties by population. This selection process identified the following affected counties:

- OR: Multnomah, Washington, Clackamas, Lane, Marion
- WA: King, Spokane, Clark, Thurston, Yakima

Next, we defined non-affected counties as those with good or moderate AQI levels during the study period. From these, we selected counties with populations in the top 25% (i.e., population size ≥ 67,976), resulting in a total of 50 unaffected counties. These were also filtered by the "Northern" indoor activity cluster to ensure that they had the same seasonal behavior of the affected counties. These unaffected counties serve as a baseline in our analyses. Table S1 compares the population sizes of affected counties with the average of non-affected counties.

| county name | state | pop |
|---|---|---|
| Washington | OR | 600689 |
| Clackamas | OR | 422160 |
| Lane | OR | 382940 |
| Marion | OR | 346202 |
| Multnomah | OR | 815871 |
| Spokane | WA | 540700 |
| King | WA | 2272571 |
| Thurston | WA | 295729 |
| Clark | WA | 505013 |
| Yakima | WA | 256533 |
| Avg. non-affected counties | | 997977 |

Table S1: Population sizes of selected counties in OR and WA state compared to average population size of non-affected counties.

### Regression Discontinuity of indoor activity seasonality

To detect discontinuities in the seasonal pattern of indoor activity, we employed the Regression Discontinuity (RD) approach. It is a statistical method used to estimate the causal effect of a treatment or intervention by leveraging a sharp discontinuity in the relationship between a continuous assignment variable and an outcome variable. In our context, the core concept of RD is based on the notion that the time-dependent indoor activity seasonality is inherently similar in the 8 weeks study period, except for the exposure to the wildfire event. By comparing the indoor seasonal activity for the affected counties at the starting date of the event, we can isolate the anomalous effect caused by wildfires from confounding factors (i.e., other potential local anomalies in the mobility).

For regression discontinuity analysis, we used a local linear regression model. It estimates the event's effect by fitting a linear regression line separately for a set of observations after and before it. The local linear regression model can be represented as follows:

$$\sigma_{it} = \lambda + \beta t_i + \gamma D_{it} + \epsilon_t,$$

where:

- $\sigma_{it}$ is the outcome variable, i.e., the indoor index seasonality for the county $i$ at the day $t$;
- $t_i$ is the temporal variable for the county i;
- $D_{it}$ is the wildfire event indicator variable, such that:

  $$D_{it} = 1 \quad \text{if} \quad t > c$$

  $$D_{it} = 0 \quad \text{if} \quad t \leq c$$

  where $c$ is the start date of the wildfire;
- $\lambda$ and $\beta$ are the coefficients representing the intercept and slope of the regression line;

- $\gamma$ is the wildfire event effect, which represents the difference in outcome between the observations before and after the event;

- $\epsilon_t$ is the error term.

To give a larger weight to observations closer to the moment of the event, we applied a triangular kernel weighting function. We quantified discontinuities in indoor activity seasonality with respect to September 2020 wildfires. Table S1 shows the results of the analysis. We observed that all affected counties in WA state show a significant increase in indoor activity seasonality after the starting date of the wildfires. This also holds for Multnomah and Clackamas counties in Oregon state. This latter is the county showing the greatest increase in indoor activity overall. The increase is not significant for Marion county in Oregon.

| state | County name | $\gamma$ | $\gamma_{5\%}$ | $\gamma_{95\%}$ |
|---|---|---|---|---|
| OR | Multnomah | 0.12 | 0.04 | 0.20 |
| OR | Washington | 0.09 | 0.02 | 0.15 |
| OR | Clackamas | 0.32 | 0.26 | 0.37 |
| OR | Lane | 0.08 | 0.02 | 0.15 |
| OR | Marion | 0.04 | -0.03 | 0.10 |
| WA | King | 0.17 | 0.13 | 0.21 |
| WA | Spokane | 0.16 | 0.11 | 0.20 |
| WA | Clark | 0.11 | 0.05 | 0.18 |
| WA | Thurston | 0.14 | 0.08 | 0.20 |
| WA | Yakima | 0.02 | 0.00 | 0.04 |

Table S1. Regression discontinuity coefficient by US county with 90% CI.

### Infectious disease model and simulation details

To characterize local infectious disease dynamics, we developed a deterministic compartmental SIR model for each county of the form:

$$\frac{dS}{dt} = -\beta_0\,\beta(t)\,\frac{SI}{N}$$

$$\frac{dI}{dt} = \beta_0\,\beta(t)\,\frac{SI}{N} - -\gamma I$$

$$\frac{dR}{dt} = \gamma I$$

where:

- $S$ represents the number of susceptible individuals;
- $I$ is the number of infectious individuals;
- $R$ is the number of recovered individuals;
- $\gamma$ is the recovery rate;
- $\beta_0\beta(t)$ is the transmissibility rate, defined as the product of a county-independent part $\beta_0$ and a county-dependent $\beta(t)$ [1]. $\beta_0$ is a constant parameter that takes into account the overall transmissibility, while $\beta(t)$ is indoor seasonality index that accounts for potential disruption of human mobility caused by wildfires;
- $N$ is the county population.

We defined the force of infection as $\lambda = \frac{I}{N}\ \beta 0\beta(t)$.

We run all the simulations starting one week before the start of the wildfire and observe the evolution of the epidemic spreading during three weeks. We compared the model's outcome for affected and unaffected counties, looking at the relative peak incidence as the relative variation of the occurrence of new cases of disease at the incidence peak day. The relative quantities are obtained by computing the relative variation between the model's outcome for affected and unaffected counties.

We explored the relative peak incidence using $R_0$ equal to 1.3, 1.5, and 3. We choose such values because 1.3 and 1.5 are compatible with a seasonal spread of respiratory viruses such as influenza and currently SARS-CoV-2, and 3.0 was close to the basic reproduction ratio of the wild-type SARS-CoV-2.

### Masking interventions

In order to model a mask intervention, we included a reduction factor m in the force of infection accounting for the people masking. We multiplied λ for a factor m = σρ + (1 – ρ) where σ is equal to 0.6, being the estimated reduction of infection attributed to mask-wearing [5], and ρ is the fraction of people wearing the mask during the wildfire. We explored several values for ρ ranging from 0.1% to 50%. We consider the masking intervention only in the days when AQI is unhealthy.

### Sensitivity analysis

We further assessed, following the same methodology, the model's outcome for affected and unaffected counties, looking at the relative variation in the attack rate as shown in Fig. S1. We observed similar patterns to the relative peak incidence analysis.

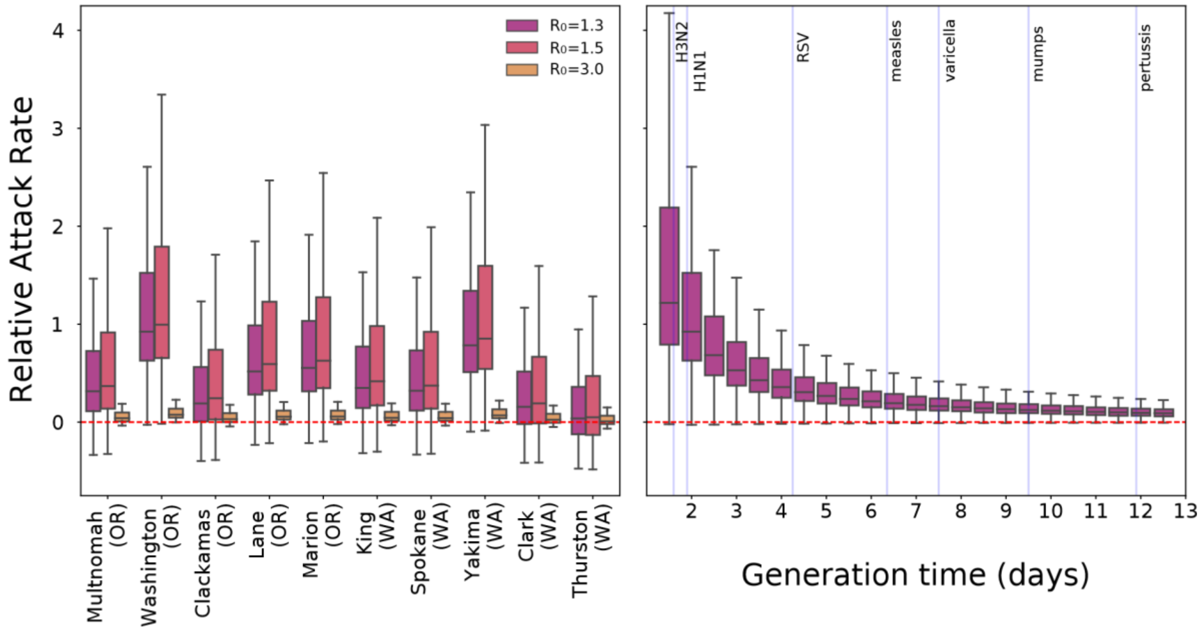


Figure S1: Left: Relative variation in attack for the selected affected counties under different infective scenarios: $R_0$ = 1.3, $R_0$ = 1.5, $R_0$ = 3.0. Right: Exploration of the relative variation in attack rate (Washington county, OR) for different values of the generation time and a fixed $R_0$ = 1.3. Blue vertical lines indicate the average generation time of different airborne respiratory diseases.
